# Overexpression of HIF-2α in Protected Regions of Alzheimer’s disease Resilient Cases

**DOI:** 10.1101/2020.08.27.20180331

**Authors:** Vivianne Mitri

## Abstract

Alzheimer’s disease (AD) Resilient individuals are characterized by having a degree of amyloid plaques at level with that of demented individuals, but a reduced amount of abnormal neurofibrillary protein “tangles” (NFTs). NFTs, also known to be upregulated under hypoxic conditions, become clinically relevant when involved in the stratum radiatum. In this paper, we show this region and more to have significant increases of hypoxic adaptive protein, HIF-2α, within AD resilient cases. Pericyte staining was present in the stratum lacunosum and radiatum of all cases affected by AD pathology (n = 4) but in AD resilient cases were increased by 12-fold (n=3) p<.0001. No staining was detected in normal cases (n=2). HIF-2α was also only present in hippocampal neuronal nuclei of AD resilient cases, including the dentate gyrus and CA1. Cytoplasmic staining of pyramidal neurons within the subiculum was seen in all cases affected by AD pathology. The intensity of HIF-2α appears to be specific to known regions of protection in AD resilience and to increase on a gradient that corresponds to protection against dementia. These results also highlight the stratum lacunosum and radiatum as regions critically impacted by hypoxic insult among AD cases.

**Significance:** HIF-2α directly regulates expression of erythropoietin (EPO), a neuroprotective glycoprotein that in brain pericytes is completely dependent upon activation of HIF-2α. To date, only indirect evidence exists that shows that brain pericyte-derived EPO can reach the bloodstream via HIF-2α expression (Urrutia et al, 2016). In this study, we provide novel preliminary findings that directly show HIF-2α expression in pericytes of human brains. Additionally, its localization is specific to the CA1 of the hippocampus, a region critical for hypoxic adaptation and the progression of Alzheimer’s disease. Finally, we present evidence of neuronal expression of HIF-2α in other critical regions of protection within AD resilient cases.

## Introduction

In clinical trials, patients diagnosed with “Alzheimer’s disease (AD)” typically exhibit dementia and brain histopathology consisting of amyloid “plaques” and neurofibrillary protein “tangles” (NFTs). However, non-demented control subjects may exhibit the same degree of histopathology, which is with the same degree of amyloid plaques but around half the amount of NFTs. The latter AD resilient patients, collectively referred to here as Non-Demented with Alzheimer’s (NDAN), deserve a more detailed investigation.

Increasing evidence suggests that hypoxia facilitates the pathogenesis of AD by increasing the hyperphosphorylation of tau and promoting the degeneration of neurons and impairing the normal functions of the blood-brain barrier (Morris, et. al 2011). Under normoxic conditions, tau is enriched in axons where it stabilizes and binds to microtubules. In AD and exposure to hypoxia, the hyperphosphorylation of tau at several serine and threonine residues reduces its ability to bind to microtubules (Morris et. al 2011). The result of this hyperphosphorylation includes destabilized microtubules and accumulation of NFTs (Schettini, 2010; Tala, 2014). This accumulation induces impaired cell division, shape changes, motility, and cell differentiation such as the formation of neuronal outgrowths. For such reasons, these features have been long regarded as the main captains in AD.

As a result of such findings, the role of hypoxia in AD is of increasing interest. However, oxygen deprivation and its relationship to NDAN has not been sufficiently investigated. Considering that NDAN individuals do not demonstrate the cognitive deficits associated with hypoxia/Alzheimer’s, it is laudable that they have managed an appropriate cellular response to hypoxic damage. This response may be responsible for reducing the NFTs in NDAN brains despite the increase in Aβ. In this paper, we show that NDAN individuals demonstrate a region-specific increase in HIF-2α, a hypoxia sensitive protein with neuroprotective factors. A further understanding of NDAN etiology guided by hypoxic adaptive mechanisms could help narrow the crucial determinants for permitting preserved cognition despite elevated levels of AD pathology.

## Materials and Methods

Paraffin-embedded human subjects of hippocampal tissue were obtained from the Sun Health Banner Research Institute in Arizona, CA. Sections were cut from 10% formalin-fixed cryoprotected tissue slabs. IHC staining and whole slide scanning were performed by NDB Bio, LLC (Baltimore, MD USA, www.ndbbio.com). A positive control for HIF-2α was confirmed using human kidney tissue, and a negative control using a section of one of the 6 human subjects (no primary antibody) was run. FFPE sections were deparaffinized and hydrated and heat-induced antigen retrieval was performed using citrate EDTA buffer. Primary Antibody used: HIF2 (Novus, cat# NB100-122SS) (dil 1:500). The reaction was developed using a biotin-free detection system and visualized using DAB. Slides were counterstained with Gill’s II hematoxylin, then dehydrated and cover slipped. All original microscope images were taken using whole slide scanning information: 20x objective (Olympus Plan N NA 0.40). File format: “.SVS”. Image J version 2.0-rc-69/1.52 p was used to analyze and compare images.

## Results

Cytoplasmic HIF-2α staining of the subiculum’s pyramidal neurons was seen only in cases affected by AD pathology, with NDAN having the strongest intensity of cytoplasmic staining, and also appearing to have the healthiest pyramidal cells in that region (Fig. 1 a & c). Size of the pyramidal neurons in this region also varied according to the level of HIF-2α expression (Fig. 6). Most notable was that all subjects affected by AD pathology had HIF-2α positive pericyte staining in the stratum lacunosum and radiatum (Fig. 2) (n=4). In AD resilient cases (n=3), staining had a 12-fold more intensity than in AD subjects (n=1) (p< 0.05) P = 0.0381. Blood vessels were also much more disfigured in this region as compared to AD resilient and control cases (Fig.2). No positive blood vessel staining for HIF-2α was detectable in normal cases (n=2). Layer II in the entorhinal cortex was also preserved among AD resilient cases and had specific staining of HIF-2α as well (Fig. 5). Other regions of CA1 in NDAN had scattered positive nuclei surrounding blood vessels but were not as uniform or intense as was staining in the stratum lacunosum (data not shown).

**Fig. 1.**
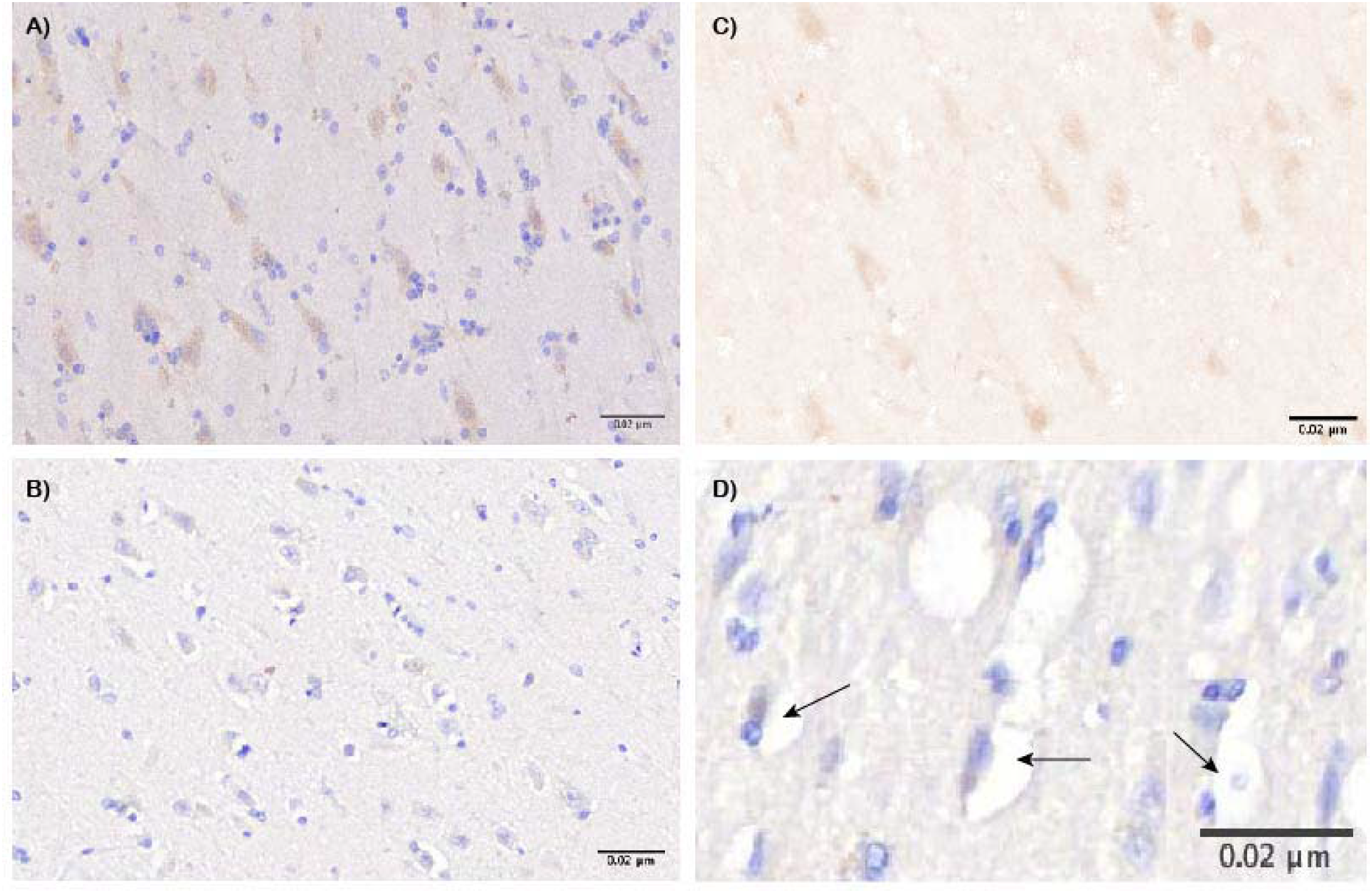
a & c Human AD resilient coronal section of subiculum taken at 20xobj. with Hematoxylin stain and HIF-2α IHC cytoplasmic staining visible by brown reaction product, b: Human coronal section of subiculum also taken at20x obj. with Hematoxylin stain and HIF-2α IHC but no visible reaction product d: Human AD subiculum with Hematoxylin stain and HIF-2α IHC taken at60xobj., and arrows pointing towards gaps where degeneration of pyramidal neurons has occured.

**Fig. 2.**
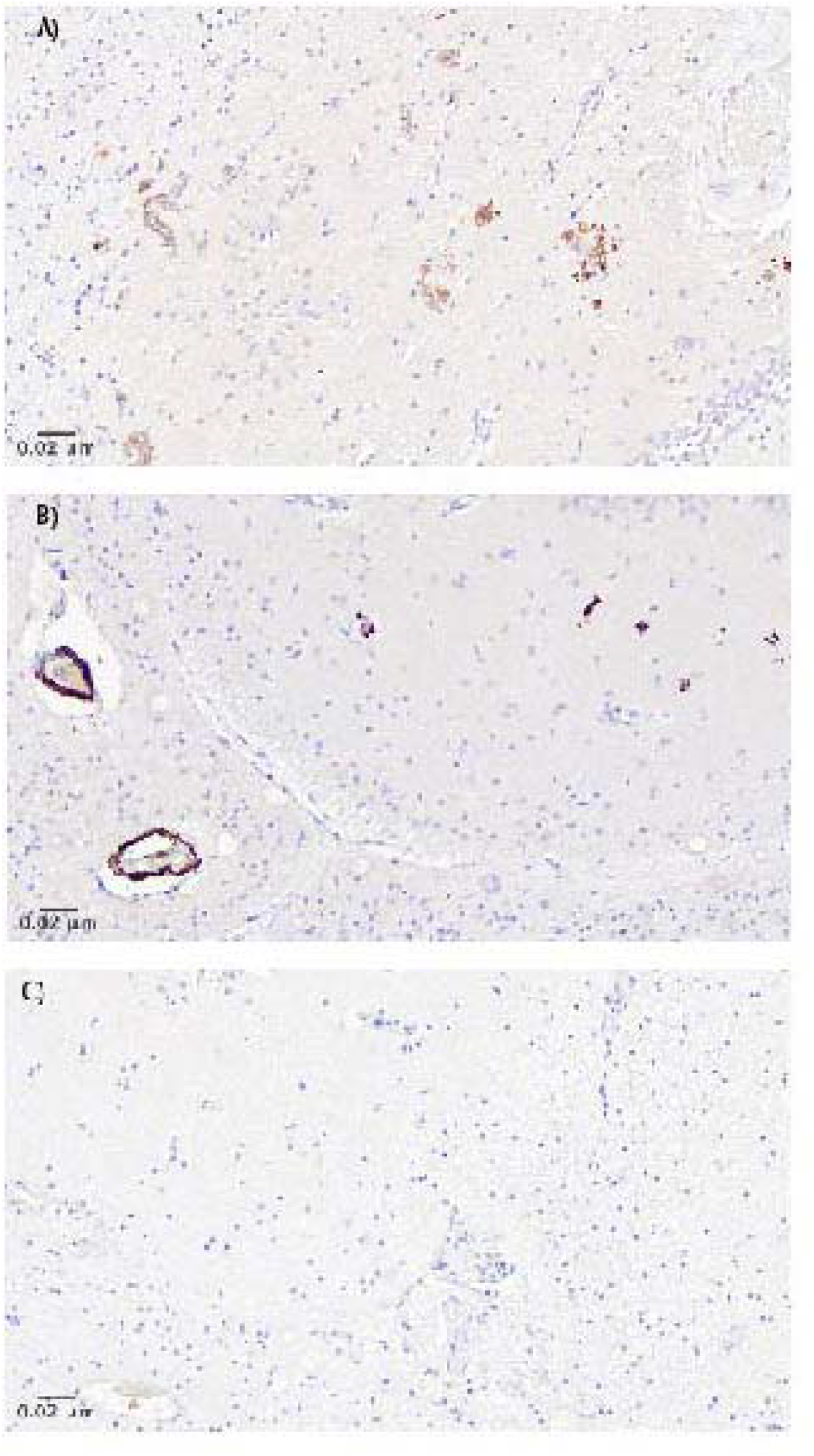
a: Coronal section of human AD subject with HIF-2α staining and disfigured pericytes with scattered HIF-2α positive nuclei in stratum lacunosum and radiatuim.Counterstained with Hematoxylin, b: Human AD resilient subject with positive HIF-2α pericytes and counter stained with Hematoxylin, c: Human control subject with lack of HIF-2α positive pericytes in stratum lacunosum and radiatum. Counter stained with Hematoxylin.

**Fig. 3.**
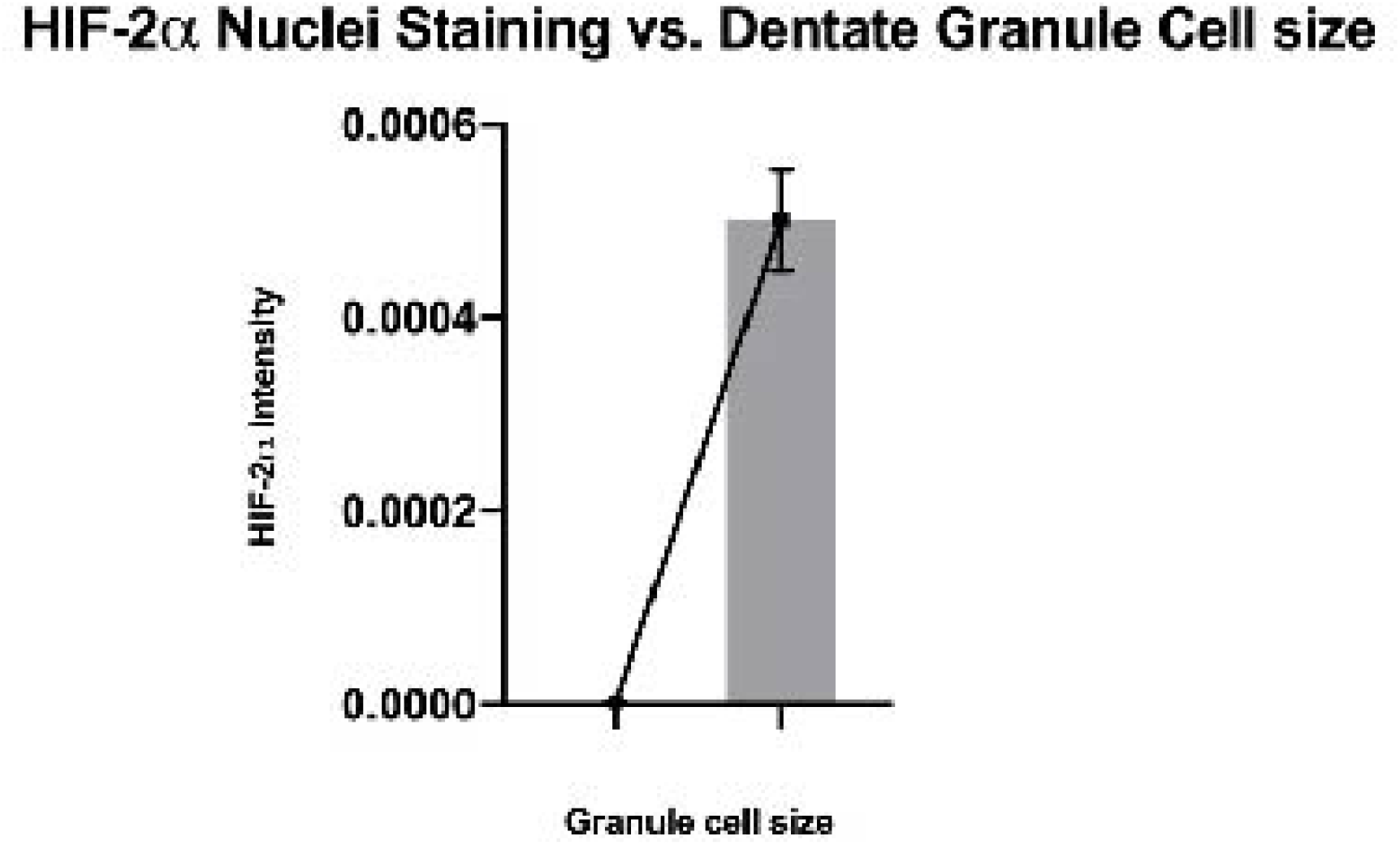
Graph depicts how dentate granule cell size varies among human subjects with AD pathology. AD resilient cases had HIF-2α positive nuclei whereas AD cases did not. n = 4. *P value (one tailed) <0.0001

**Fig. 4.**
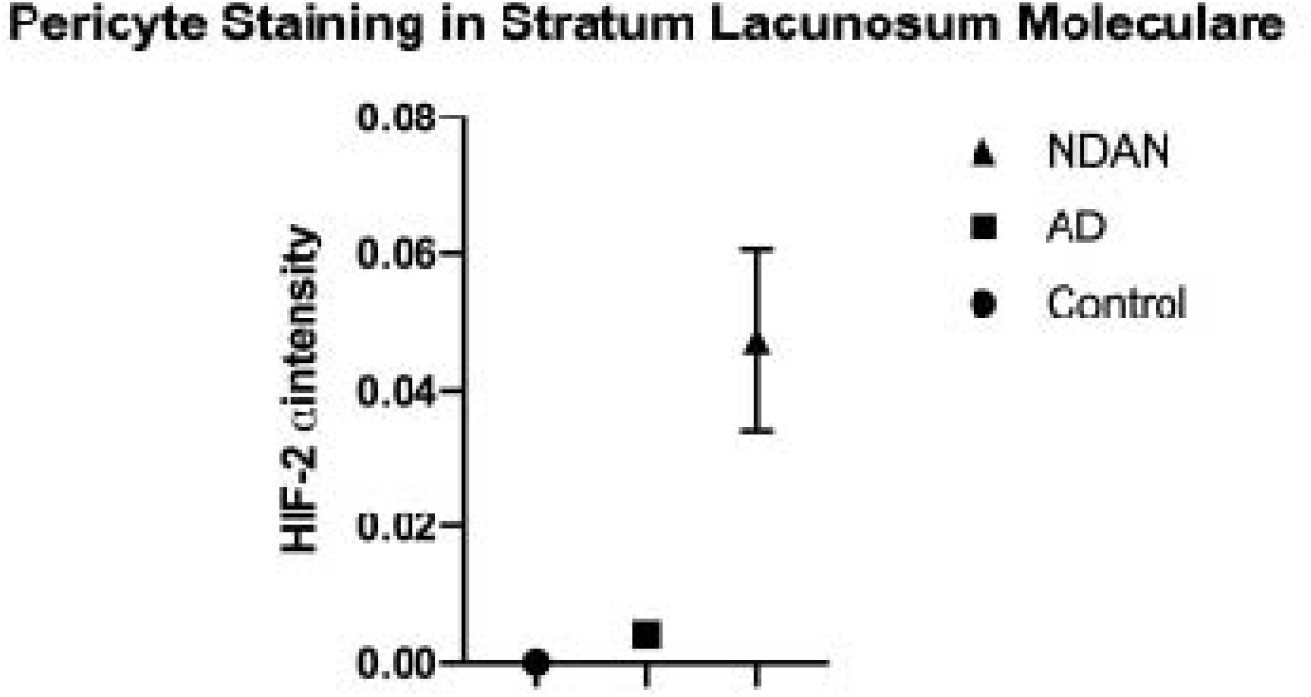
Graph shows varying levels of HIF-2α expression in the pericytes of stratum lacunosum among human AD resilient (NDANJ, AD and control subjects with SEM. NDAN cases had an average 12 fold increase in HIF-2α expression in pericytes as com pared to AD, whereas control cases had no detectable staining (p<0.05).

**Fig. 5.**
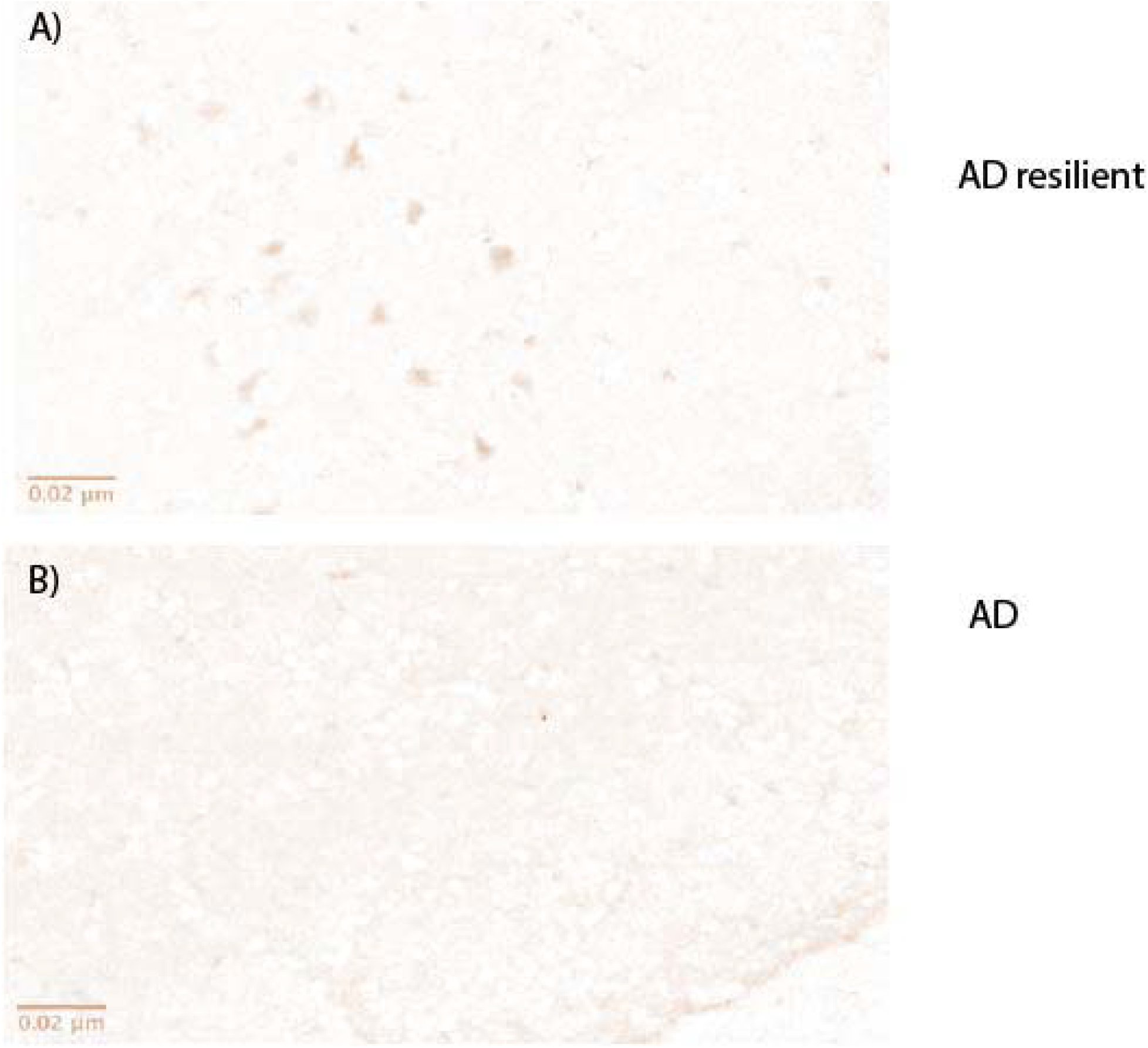
a: Shows coronal section of human subject with HIF-2α positive layer II entorhinal cortex (EC) cells, also known to be protected in NDAN (AD resilient) cases. Image J Color Deconvoluter was used to separate DAB from Hematoxylin stain. b: shows coronal section of human subject with AD and no visible staining of HIF-2α Image J Color Deconvoluter was used to separate DAB from Hematoxylin stain

**Fig. 6.**
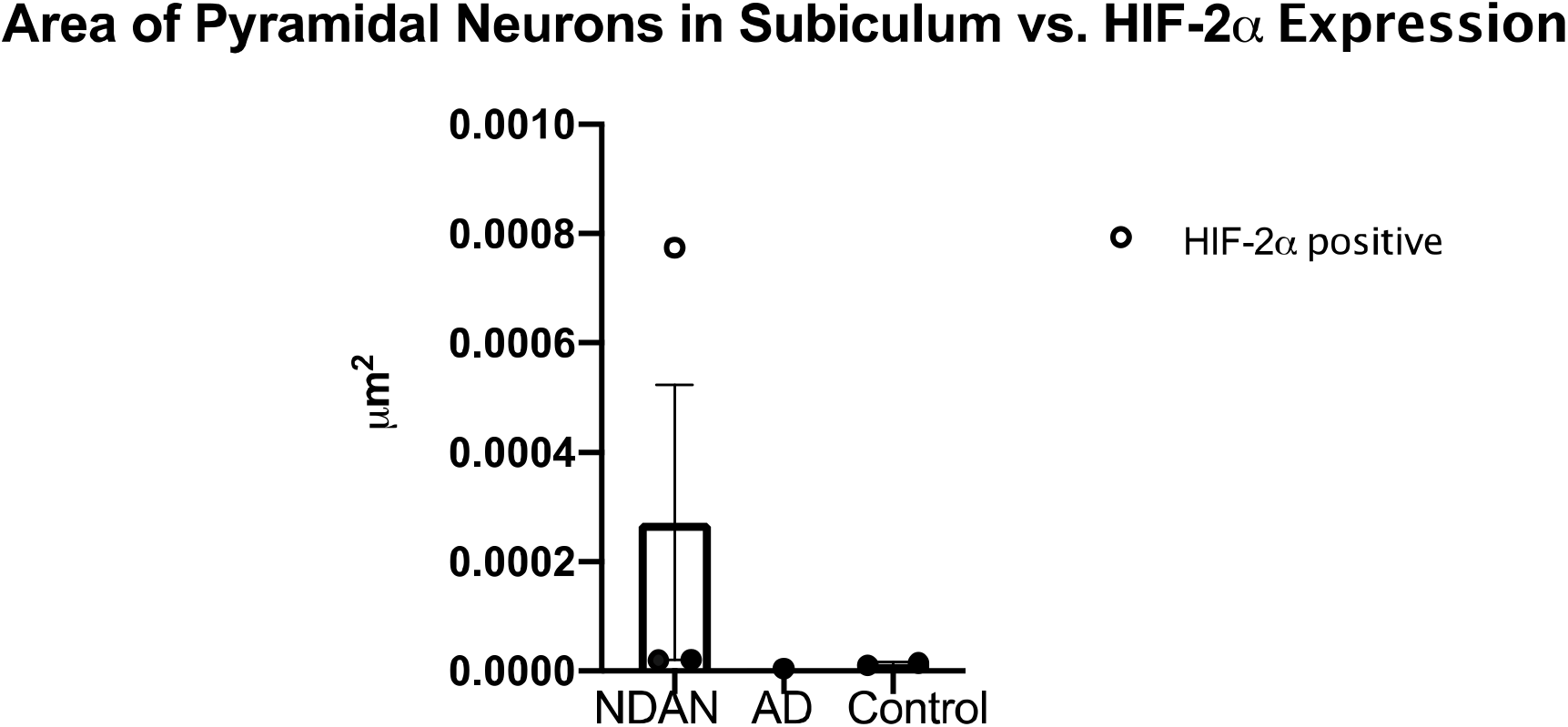
Shows pyramidal neuron size difference among groups. Intensity of HIF-2α expression was localized to the cytoplasm of pyramidal neurons in the subiculum and level of intensity was confirmed by an objective neuropathologist. Staining was only seen in NDAN (AD resilient) and AD groups and intensity varied from “positive” to “weak/mild”. Only one case showed a strong “positive” result. There was no visible expression in control group.

**Table 1.** Data Set Table on Cases studied with varying levels of Alzheimer’s pathology and Test Scores.

| CaselD | expired_age | _mmse_test_sc | PMI | MCI | ApoE | PlaqueTotal | Plaque density | TangleTotal | Braak score | neurological_dx |
| --- | --- | --- | --- | --- | --- | --- | --- | --- | --- | --- |
| AD 97-21 | 81 |  | 1.75 | no | 3/4 | 13 | frequent | 15 | VI | Alzheimer's disease |
| Control 03-41 | 89 | 29 | 2.5 | no | 3/4 | 5.25 | moderate | 1 | II | Control |
| Control 03-12 | 88 | 29 | 7 | no | 3/4 | 0 | zero | 3 | III | Control |
| NDAN 14-05 | 92 | 30 | 2.87 | no | 3/3 | 9 | frequent | 6 | IV | Control |
| NDAN 09-44 | 97 | 28 | 3.5 | no | 3/3 | 12.5 | frequent | 5 | IV | Control |
| NDAN 11-102 | 93 | 25 | 2.08 | no | 2/3 | 10 | frequent | 6 | IV | Control |

## Discussion

AD resilience is a phenomenon that is gaining attention in the research field as one that is more intricate than just “preclinical” AD. In this study, we showed that the level of HIF-2α has region-specific increases only in AD affected human subjects and that the most commonplace for its strong expression is in the pericytes of the stratum lacunosum and radiatum. Based on our results, this area appears to be a losing battle in AD in terms of HIF-2α response when compared to AD resilient cases.

All regions affected by HIF-2α in this study are mostly known to be protected in NDAN but are also critical for the progression of tau hyperphosphorylation, such as the CA1/subiculum border, dentate gyrus and most notably, the stratum lacunosum and radiatum (Thal et al, 2000; Lace et al, 2009). Several other regions affected by HIF-2α did not have a uniform level of expression. For example, some NDAN cases showed neurons with positive nuclei staining in CA1 while the other two NDAN cases had more cytoplasmic staining in that region. That being said, only NDAN cases had positive hippocampal neurons that stained for HIF-2α, and the finding that layer II cells of EC were both preserved and HIF-2α positive may help explain why NDAN cases have been reported to have the protection of layer II cells (Arnold, *et al* 2013). This has always been a perplexing find, though with these results now made clearer, considering that layer II of the EC is significantly compromised in very early stages of AD pathology (Gomez-Isla T et al, 1996). These findings, namely the expression of HIF-2α in the nuclei CA1 neurons, also help shed light onto why the nuclei in this region have been reported to not only have unique protection but also significant hypertrophy in NDAN cases as compared to both AD and control subjects (Iacono *et al*, 2009).

Among all three groups, cytoplasmic staining of the pyramidal neurons in the subiculum was only seen in those affected by AD pathology. The size of those cells, namely their dendritic length, appeared to increase in length with HIF-2α cytoplasmic intensity. More studies done with antibodies specific to neurons and dendritic analysis would help us to objectively measure the difference in dendrite length among groups and to compare results with varying levels of HIF-2α expression.

## Conclusion

In this study, the most significant difference in HIF-2α expression was visible in the pericytes of all human subjects affected by AD pathology, with a 12-fold increase in expression of HIF-2α detected in AD resilient cases. Pericytes are known to serve as oxygen sensors in the brain and are responsible for secreting the protective factor and HIF-2α regulated hormone EPO in response to hypoxic insult (Ji, 2016). This study’s most clear finding is that the stratum lacunosum and radiatum appears to be the primary region of hypoxic insult in AD. In spite of the obvious limitations of our study, this is the first report of such significant levels of HIF-2α in brains of human subjects with AD pathology.

## Data Availability

Not applicable

## References

1. Andrade-Moraes CH, Oliveira-Pinto AV, Castro-Fonseca E et al. (2013) Cell number changes in Alzheimer’s disease relate to dementia, not to plaques and tangles. Brain; 136:3738–3752. doi:10.1093/brain/awt273

2. Arnold, Steven E., et al. “Cellular, synaptic, and biochemical features of resilient cognition in Alzheimer’s disease.” Neurobiology of aging 34.1 (2013): 157-168.

3. Atwood CS, Huang X, Moir RD, Tanzi RE, Bush AI. (1999) Role of free radicals and metal ions in the pathogenesis of Alzheimer’s disease. Met Ions Biol Syst.;36:309–364.

4. Beatriz G. Perez-Nievas, Thor D. Stein, Hwan-Ching Tai, Oriol Dols-Icardo, Thomas C. Scotton, Isabel Barroeta-Espar, Leticia Fernandez-Carballo, Estibaliz Lopez de Munain, Jesus Perez, Marta Marquie, Alberto Serrano-Pozo, Mathew P. Frosch, Val Lowe, Joseph E. Parisi, Ronald C. Petersen, Milos D. Ikonomovic, Oscar L. López, William Klunk, Bradley T. Hyman, Teresa Gómez-Isla; (2013) Dissecting phenotypic traits linked to human resilience to Alzheimer’s pathology. Brain; 136 (8): 2510-2526. doi: 10.1093/brain/awt171

5. Benes FM, Sorensen I, Bird ED. (1991) Reduced neuronal size in posterior hippocampus of schizophrenic patients. Schizophrenia Bulletin;17, 597–608.

6. Berlau DJ, Corrada MM, Head E, et al. (2009) apoE epsilon2 is associated with intact cognition but increased Alzheimer pathology in the oldest old. Neurology.;72:829–834.

7. Boutet, Claire, et al. “Detection of volume loss in hippocampal layers in Alzheimer’s disease using 7 T MRI: a feasibility study.” NeuroImage: Clinical 5 (2014): 341-348.

8. Braak, E., and H. Braak. “Alzheimer’s disease: transiently developing dendritic changes in pyramidal cells of sector CA1 of the Ammon’s horn.” Acta neuropathologica 93.4 (1997): 323-325.

9. Bush AI. 2003 The metallobiology of Alzheimer’s disease. Trends Neurosci.;26:207–214

10. Björklund NL, Reese LC, Sadagoparamanujam VM, Ghirardi V, Woltjer RL, Taglialatela G (2012) Absence of amyloid β oligomers at the postsynapse and regulated synaptic Zn2+ in cognitively intact aged individuals with Alzheimer’s disease neuropathology. Mol Neurodegener;7:23 pmid:22640423

11. Cai XD, Golde TE and Younkin SG. (1993) Release of excess amyloid β protein from a mutant amyloid β protein precursor. Science; 259, 514–516.

12. Castellani RJ, Lee HG, Zhu X, et al. (2006) Neuropathology of Alzheimer disease: pathognomonic but not pathogenic. Acta Neuropathol (Berl); 111:503–9

13. Cavadas, MAS. et al. (2016) REST is a hypoxia-responsive transcriptional repressor. Sci. Rep.; 6, 31355; doi: 10.1038/srep31355.

14. Chen, J., Gu, Z., Wu, M., Yang, Y., Zhang, J., Ou, J., … Chen, Y. (2016). C-reactive protein can upregulate VEGF expression to promote ADSC-induced angiogenesis by activating HIF-1α via CD64/PI3k/Akt and MAPK/ERK signaling pathways. Stem Cell Research & Therapy, 7, 114. http://doi.org/10.1186/s13287-016-0377-1

15. Chepelev, NL, Bennitz, JD, Huang, T, McBride, S. & Willmore, WG (2011) The Nrf1 CNC-bZIP protein is regulated by the proteasome and activated by hypoxia. PLoS One 6, e29167.

16. Cipolla MJ (2010) The cerebral circulation. In: Integrated Systems Physiology: From Molecule to Function, (Granger DN, Granger J. San Rafael ed), pp1–59. Morgan & Claypool Life Sciences.

17. Citron M, Oltersdorf T, Haass C, McConlogue L, Hung A. Y., Seubert P, Vigo-Pelfrey C, Lieberburg I, Selkoe DJ (1992) Mutation of the beta-amyloid precursor protein in familial Alzheimer’s disease increases β-protein production. Nature;360, 672–674.

18. Cooper GM (2000) The Cell: A Molecular Approach. 2nd edition. Sunderland (MA): Sinauer Associates.

19. Codispoti KE et al. (2012) Longitudinal brain activity changes in asymptomatic Alzheimer disease. Brain Behav. 2(3):221–230.

20. Cotter, D, Mackay, D, Landau, S, Kerwin, R & Everall, I (2001) Reduced glial cell density and neuronal size in the anterior cingulate cortex in major depressive disorder. Archives of General Psychiatry, 58, 545–553.

21. Dickson DW, Crystal HA, Mattiace LA, Masur DM, Blau AD, Davies P, Yen SH, Aronson MK (1992) Identification of normal and pathological aging in prospectively studied nondemented elderly humans. Neurobiol Aging; 13: 179–189.

22. Driscoll and Troncoso, Driscoll I, Troncoso J (2011) Asymptomatic Alzheimer’s disease: a prodrome or a state of resilience? Curr. Alzheimer Res.; 8 pp. 330–335

23. Dumont M, Stack, C., Elipenahli, C., Jainuddin, S, Launay, N, Gerges, M,. Pujol, A. (2014). PGC-1α overexpression exacerbates β-amyloid and tau deposition in a transgenic mouse model of Alzheimer’s disease. The FASEB Journal, 28(4), 1745–1755.

24. Dweik RA (2005) Nitric oxide, hypoxia, and superoxide: the good, the bad, and the ugly! Thorax, 60:265–267

25. Ferrara N, Gerber, HP, LeCouter, J. (2003). The biology of VEGF and its receptors. Nature medicine, 9(6), 669–676.

26. da Fonseca RR, Johnson WE, O’Brien SJ, Ramos MJ, Antunes A (2008) The adaptive evolution of the mammalian mitochondrial genome. BMC Genomics.; 9:119

27. Frederickson, CJ, Rampy, BA, Reamy-Rampy, S., Howell, GA (1992) Distribution of histochemically reactive zinc in the forebrain of the rat. J. Chem. Neuroanat. 5: 521–530.

28. Galbán, CJ et al. 2009 The parametric response map is an imaging biomarker for early cancer treatment outcome. Nat.. Med. 15, 572–576

29. Gannon, Mary et al. Noradrenergic dysfunction in Alzheimer’s disease. (2015) Frontiers in Neuroscience.; Vol. 9 p1-12. 12p.

30. Giannakopoulos P, Herrmann FR, Bussiere T, Bouras C, Kovari E, Perl DP, Morrison JH, Gold G, Hof PR (2008) Tangle and neuron numbers, but not amyloid load, predict cognitive status in Alzheimer’s disease. Neurology. 60:1495–1500

31. Gour, N. et al. (2008) Functional connectivity changes differ in early and late-onset Alzheimer’s disease. Hum. Brain Mapp.; 35, 2978–2994.

32. Gutsaeva DR, Carraway MS, Suliman HB, Demchenko IT, Shitara H, Yonekawa H, and Piantadosi CA. (2008) Transient hypoxia stimulates mitochondrial biogenesis in brain subcortex by a neuronal nitric oxide synthase-dependent mechanism. J Neurosci. 28: 2015–2024

33. Hara, H., Wada, T., Bakal, C., Kozieradzki, I., Suzuki, S., Suzuki, N., … & D’acquisto, F. (2003). The MAGUK family protein CARD11 is essential for lymphocyte activation. Immunity, 18(6), 763–775.

34. Head E, Corrada MM, Kahle-Wrobleski K, et al. (2009) Synaptic proteins, neuropathology and cognitive status in the oldest-old. Neurobiol Aging;30:1125–1134.

35. Hoffman, WE, Albrecht, RF, Miletich, DJ (1984). The role of adenosine in CBF increases during hypoxia in young vs aged rats. Stroke, 15(1), 124–129.

36. Holmes C, Boche D, Wilkinson D (2008) et al. Long-term effects of Abeta42 immunisation in Alzheimer’s disease: follow-up of a randomised, placebo-controlled phase I trial. Lancet 372, 216–223

37. Hill BG, Benavides GA, Lancaster JR, et al. (2012) Integration of cellular bioenergetics with mitochondrial quality control and autophagy. Biological chemistry.;393(12):1485–1512. doi:10.1515/hsz-2012-0198.

38. Iacono D, Markesbery WR, Gross M et al. (2009) The Nun study: clinically silent AD, neuronal hypertrophy, and linguistic skills in early life. Neurology; 73: 665–73.

39. Jackson DC Acid-base balance during hypoxic hypometabolism: selected vertebrate strategies. Respir. Physiol. Neurobiol. 2014; 141, 273–283

40. Jonsson, SA, Luts, A, Guldberg-Kjaer N, Ohman, R (1999) Pyramidal neuron size in the hippocampus of schizophrenics correlates with total cell count and degree of cell disarray. European Archives of Psychiatry and Clinical Neuroscience, 249, 169–173.

41. Jensen, S, Gassama, MP, Heidmann, T. Cosuppression of I transposon activity in Drosophila by I-containing sense and antisense transgenes. Genetics (1999) 153(4): 1767--1774

42. Kokjohn TA, Maarouf CL, Roher AE. (2012) Is Alzheimer’s disease amyloidosis the result of a repair mechanism gone astray? Alzheimer’s & dementia: the journal of the Alzheimer’s Association;8(6):574–583. doi:10.1016/j.jalz.2011.05.2429.

43. Kumar-Singh S., Theuns J., Van Broeck B., Pirici D., Vennekens K., Corsmit E., Cruts M., Dermaut B., Wang R. and Van Broeckhoven C. (2006) Mean age-of-onset of familial alzheimer disease caused by presenilin mutations correlates with both increased Abeta42 and decreased Abeta40. Hum Mutat. 27, 686–695.

44. Lee B, Butcher GQ, Hoyt KR, Impey S., Obrietan K (2005) Activity-dependent neuroprotection and cAMP response element-binding protein (CREB): kinase coupling, stimulus intensity, and temporal regulation of CREB phosphorylation at serine 133. J. Neurosci.; 25, 1137–1148.

45. Li RC, Haribabu B, Mathis SP, Kim, J and Gozal D (2011) Leukotriene B4 Receptor-1 mediates Intermittent Hypoxia-induced Atherogenesis. Am J Respir Crit Care Med 184(1): 124–31.

46. Lih-Fen L, Libuse B, Harold WC, Joseph R. (1996) Inflammation, Aβ Deposition, and Neurofibrillary Tangle Formation as Correlates of Alzheimer’s Disease Neurodegeneration. J Neuropathol Exp Neurol.; 55 (10): 1083-1088. doi: 10.1097/00005072-199655100-00008

47. Lu, T., Aron, L., Zullo, J., Pan, Y., Kim, H., Chen, Y., Bennett, D. A. (2014). REST and stress resistance in ageing and Alzheimer’s disease. Nature, 507(7493), 448.

48. Marrif, H. and Juurlink, BH. (1999) Astrocytes respond to hypoxia by increasing glycolytic capacity. J. Neurosci. Res. 57: 255–260.

49. Martínez, M., Hernández, A. I., & Hernanz, A. (2001). Increased cAMP immunostaining in cerebral vessels in Alzheimer’s disease. Brain research, 922(1), 148–152.

50. Morris M, Maeda S, Vossel K, Mucke L. (2011) The Many Faces of Tau. Neuron.;70(3): 410-426. doi:10.1016/j.neuron.2011.04.009.

51. von Gunten A, et al. (2010) Brain aging in the oldest-old. Curr Gerontol Geriatr Res. Epub.

52. Moussavi Nik SH., Wilson L, Newman M, Croft K, Mori TA, Musgrave I, et al. (2012) The BACE1-PSEN-AbetaPP regulatory axis has an ancient role in response to low oxygen/oxidative stress. J. Alzheimers Dis. 28 515-530 10.3233/JAD-2011-110533

53. Nilsson K, Gustafson L, Hultberg B. C-reactive protein level is decreased in patients with Alzheimer’s disease and related to cognitive function and survival time. Clin Biochem. (2011);44(14-15):1205–1208

54. Norris, ML, and Millhorn, DE J. Biol. Chem. 1995; 270, 23774–23779

55. Norris, ML, and Millhorn, DE J. Biol. Chem. 1995 270, 23774–23779

56. Oberlander, JG, Schlinger, BA, Clayton, NS, & Saldanha, CJ Neural Aromatization accelerates the acquisition of spatial memory via an influence on songbird hippocampus. Horm Behav. 2004; 45 250–258

57. Padurariu, M, Ciobica, A, Mavroudis, I, Fotiou, D, and Baloyannis, S. Hippocampal neuronal loss in the CA1 and CA3 areas of Alzheimer’s disease patients. Psychiatr. Danub. 2012 24, 152–158.

58. Ross JM, Oberg J, Brene S, Coppotelli G, Terzioglu M, Pernold K, Goiny M, Sitnikov R, Kehr J, Trifunovic A, Larsson NG, Hoffer BJ, Olson L. (2012) High brain lactate is a hallmark of aging and caused by a shift in the lactate dehydrogenase A/B ratio. Proc Natl Acad Sci USA. 107(46):20087–20092. doi:10.1073/pnas.1008189107

59. Schettini G, Govoni S, Racchi M, Rodriguez G. Phosphorylation of APP-CTF-AICD. Journal of Neurochem. (2010) 115(6): 1299–308. doi: 10.1111/j.1471-4159.2010.07044

60. Sharp, FR Bernaudin, M. (2004). HIF1 and oxygen sensing in the brain. Nature Reviews Neuroscience, 5(6), 437–448.

61. Spillantini MG, Goedert M. Tau protein pathology in neurodegenerative diseases.(1998) Trends Neurosci;21:428–433.

62. Spires-Jones, TL, Hyman, B. T. (2014). The intersection of amyloid beta and tau at synapses in Alzheimer’s disease. Neuron, 82(4), 756–771.

63. Stockmeier, C.A., Mahajan, G.J., Overholser, J.C., Jurjus, G.J., Meltzer, H.Y., Uylings, H.B.M., Friedman, L. & Rajkawska, G. (2004) Cellular Changes in the postmortem hippocampus in major depression. Society of Biological Psychiatry.56, 640–650.

64. Tala, Sun X, Chen J, Zhang L, Liu N, Zhou J, et al. (2014) Microtubule Stabilization by Mdp3 Is Partially Attributed to Its Modulation of HDAC6 in Addition to Its Association with Tubulin and Microtubules. PLoS ONE. 9(3): e90932. doi:10.1371/journal.pone.0090932

65. Tanaka, K. (2001). Alteration of second messengers during acute cerebral ischemia–adenylate cyclase, cyclic AMP-dependent protein kinase, and cyclic AMP response element binding protein. Progress in neurobiology, 65(2), 173–207.

66. Thakur A, Wang X, Siedlak SL, Perry G, Smith MA, (2007) et al. c-Jun phosphorylation in Alzheimer disease. J Neurosci Res 85(8): 1668–73.

67. Tsui AKY, Marsden PA, Mazer CD, et al. (2011) Priming of hypoxia-inducible factor by neuronal nitric oxide synthase is essential for adaptive responses to severe anemia. Proceedings of the National Academy of Sciences of the United States of America.;108(42):17544–17549. doi:10.1073/pnas.1114026108.

68. Vangeison G, Carr D, Federoff HJ, Rempe DA. (2008) The good, the bad, and the cell type-specific roles of hypoxia inducible factor-1α in neurons and astrocytes. J. Neurosci. 28, 1988–1993.

69. Von Gunten A, Ebbing K, Imhof A, Giannakopoulos P, Kovari E (2010) Brain aging in the oldest-old. Curr Gerontol Geriatr Res 2010: 358531.

70. Wei, Caihong, et al. “Genome-wide analysis reveals adaptation to high altitudes in Tibetan sheep.” Scientific reports 6 (2016): 26770.

71. Yarchoan M, Xie SX, Kling MA, et al. Cerebrovascular atherosclerosis correlates with Alzheimer pathology in neurodegenerative dementias (2012) Brain;135(Pt 12):3749-3756

72. Yoon D, Pastore YD, Divoky V, Liu E, Mlodnicka AE, Rainey K, Ponka P, Semenza G. L, Schumacher A, Prchal JT. Hypoxia-inducible factor-1 deficiency results in dysregulated erythropoiesis signaling and iron homeostasis in mouse development. J. Biol. Chem. (2006) 281, 25703–25711

73. Zhang J, Haddad GG, Xia Y. (2006) Delta-, but not mu- and kappa-, opioid receptor activation protects neocortical neurons from glutamate-induced excitotoxic injury. Brain. Res 885:143–153.

74. Zhang X, Zhou K, Wang R, Cui J, Lipton SA, Liao FF, Xu H, Zhang Y. (2007) Hypoxia-inducible factor 1alpha (HIF-1alpha)-mediated hypoxia increases BACE1 expression and beta-amyloid generation. J Biol Chem. 282 (15): 10873-10880. 10.1074/jbc.M608856200.

75. Zhu X, Perry G, Moreira PI, Aliev G, Cash AD, Hirai K, Smith MA. J, (2006) Alzheimer’s Dis.;9:147.

76. Ziello, JE, Jovin, IS, Huang, Y. (2007). Hypoxia-Inducible Factor (HIF)-1 Regulatory Pathway and its Potential for Therapeutic Intervention in Malignancy and Ischemia. The Yale Journal of Biology and Medicine, 80(2), 51–60.

77. Zou, J., & Crews, F. (2006). CREB and NF-κB transcription factors regulate sensitivity to excitotoxic and oxidative stress induced neuronal cell death. Cellular and molecular neurobiology, 26(4-6), 383-403.

78. Xiang Y, Xu G, Weigel-Van Aken KAK, (2006). Lactic Acid Induces Aberrant Amyloid Precursor Protein Processing by Promoting Its Interaction with Endoplasmic Reticulum Chaperone Proteins. PLoS ONE; 5(11): e13820. doi:10.1371/journal.pone.0013820

